# AveloMask, a novel breath aerosol collection kit for airborne *Mycobacterium tuberculosis*: a proof-of-principle assessment

**DOI:** 10.1101/2025.04.10.25325579

**Authors:** Patricia Risch, Tobias Broger, Zandile Booi, Katie Tiseo, Harshitha Santhosh Kumar, Jamie van Schalkwyk, Reto Willi, Stefan M Botha, Peter Sander, Adithya Cattamanchi, Claudia M Denkinger, Grant Theron, Christina Fialová, Christian Adlhart, Rouxjeane Venter

**Author notes:** Contributed equally (Patricia Risch, Tobias Broger). Contributed equally (Christian Adlhart, Rouxjeane Venter). Corresponding author: Tobias Broger, Avelo Inc., Rütistrasse 16, CH-8952 Schlieren, Switzerland.

## Abstract

**Background:** Tuberculosis (TB) remains the world’s deadliest infectious disease, with sputum-based diagnostics failing to detect many active cases, often due to difficulty in specimen collection. Breath aerosols, a major route of *Mycobacterium tuberculosis* (MTB) transmission, offer a promising non-invasive alternative. This study evaluated the diagnostic accuracy and feasibility of the AveloMask, a novel breath aerosol collection kit, designed for point-of-care collection, for detecting active pulmonary TB using PCR.

**Methods:** We conducted a diagnostic accuracy study among adult outpatients with TB symptoms attending primary healthcare facilities in Cape Town, South Africa. Participants wore the mask for 45 minutes, coughing deeply five times at the start and end of collection. Breath aerosol samples were collected on a fiber filter integrated into the mask, immediately stabilized in buffer post-collection, biobanked, and later analysed by quantitative PCR (qPCR) targeting the MTB-specific *IS6110* insertion sequence. Diagnostic accuracy was assessed against sputum Xpert MTB/RIF Ultra (SXRS) and a composite microbiological reference standard (MRS), including culture. Usability was evaluated using structured questionnaires.

**Results:** Of 61 participants enrolled, 58 provided evaluable breath samples and 59% (34/58) had confirmed TB. Compared with the SXRS, mask qPCR achieved a sensitivity of 71.0% (95% CI: 53.4–83.9%) and specificity of 92.3% (95% CI: 75.9–97.9%). Compared with the MRS, sensitivity was 64.7% (95% CI: 47.9–78.5%) and specificity 91.7% (95% CI: 74.2–97.7%). Mask qPCR positivity rates increased with higher sputum bacterial loads, reaching 100% sensitivity among participants with high sputum MTB concentrations. MTB IS6110 copy numbers in extracted mask samples varied widely (range: 4–2147 copies; mean: 175 copies), but were low overall, likely reflecting incomplete DNA recovery during lysis or extraction and/or a low number of MTB bacilli in breath aerosols. Usability feedback showed that the mask and collection procedure were well-tolerated.

**Conclusions:** The AveloMask breath aerosol sampling kit demonstrated promising diagnostic accuracy for active TB, comparable to other mask-based methods, while offering ease-of-use and feasibility at the point-of-care. Future studies should improve lysis and extraction and explore integration with commercial molecular diagnostic platforms, validate these findings in larger, more diverse populations, and for different use-cases.

## Background/Introduction

Worldwide, tuberculosis (TB) reclaimed its position as the deadliest infectious disease in 2023, claiming 1.25 If Sustainable Development Goal (SDG) targets are not met, TB is projected to cause 31.8 million deaths and economic losses of US$17.5 trillion over the next 30 years.^2^ Currently, 25% of TB cases (2.7/10.8 million) go undiagnosed.^1^ A major contributor to this diagnostic gap is that 7.6% to 18% of individuals with TB are unable to produce sputum, the primary specimen used for bacteriological detection of *Mycobacterium tuberculosis* (MTB).^3–5^ Testing on alternative non-sputum-based samples has been identified as an important priority to increase diagnostic yield and help close the diagnostic gap.^6,7^

Since antiquity, it has been known that breath contains clues to many diseases.^8^ >Breath is an attractive sample type due to its non-invasive and simple collection process. In recent years, breath-based diagnostic research has primarily focused on volatile organic compounds (VOCs), which originate from metabolic processes. However, VOC-based diagnostics often lack specificity, as VOC profiles can be influenced by multiple diseases, physiological conditions, and host factors. A recent meta-analysis found a pooled specificity of 83% for VOC-based TB detection, with high heterogeneity.^9^

An alternative approach is the collection and testing of exhaled breath aerosols (XBA). Aerosols are microscopic liquid and solid particles with a size between approx. 10 nm and 100 µm that can carry pathogens, nucleic acids, and proteins.^10^ Aerosol collection and testing has the potential for higher specificity than VOC-based testing, as it enables direct detection of pathogen markers such as nucleic acids with molecular tests. Moreover, because breath aerosols are linked to respiratory pathogen transmission, XBA-based detection could help identify individuals most likely to spread infection.^11^ Growing evidence supports aerosol transmission as a major route for many respiratory infections, including pandemic coronaviruses, influenza, respiratory syncytial virus (RSV), and TB.^10,12–16^ Thus far, aerosol collection for pathogen detection was primarily done in academic research labs using techniques such as impingers, impactors, or precipitators which require relatively high technical efforts.^12,17–19^ For diagnosis of active TB, others have used face mask sampling (FMS) with PCR detection and have reported sensitivities ranging from 65% to 86%.^20–23^ >An important operational consideration for FMS is the removal and processing of the aerosol capture filter for subsequent testing, which is typically done aseptically with forceps, prone to contamination, and often impractical at the point of collection. Other key aspects are the highly efficient aerosol capture and release combined with compatibility of the recovered aerosol sample with downstream extraction, molecular testing, and sample transport stability. To address these challenges, we have developed an easy-to-use FMS kit (AveloMask Kit, Avelo, Switzerland) for the collection, concentration, transport, and storage of human breath aerosol specimens.

In this study, we aimed to assess the feasibility and diagnostic accuracy of AveloMask collection combined with quantitative polymerase chain reaction (qPCR) for the detection of active TB in adult outpatients, compared against a composite microbiological reference standard (MRS) and sputum Xpert MTB/RIF Ultra (Xpert Ultra; Cepheid). In addition, we aimed to quantify MTB *IS6110* copies recovered from mask samples and to evaluate losses due to incomplete lysis to inform further protocol improvements.

## Methods

### AveloMask kit design and procedure

AveloMask (Avelo AG, Switzerland) is a specimen collection kit for the collection of a person’s breath sample from the respiratory tract. The processing steps are illustrated in Figure 1. The kit contains a mask and a buffer tube. The inner side of the mask contains a filter inlay to collect aerosol particles from the exhalate. The filter inlays are 75 x75 mm sheets with electrospun fibers mounted to the inside of the mask as described in the supplemental material. After wearing the mask (step 1), two stickers from each side of the mask, which protect the inlay during wear, are peeled off (step 2), and the filter inlay is removed by pushing it into the buffer tube using a stick attached to the tube cap (step 3). The buffer inactivates the sample and preserves nucleic acids for transport of the sample at ambient temperature until further processing or biobanking (step 4). Detailed instructions are given in Figure S1 and the video in the supplemental material.

**Figure 1:**
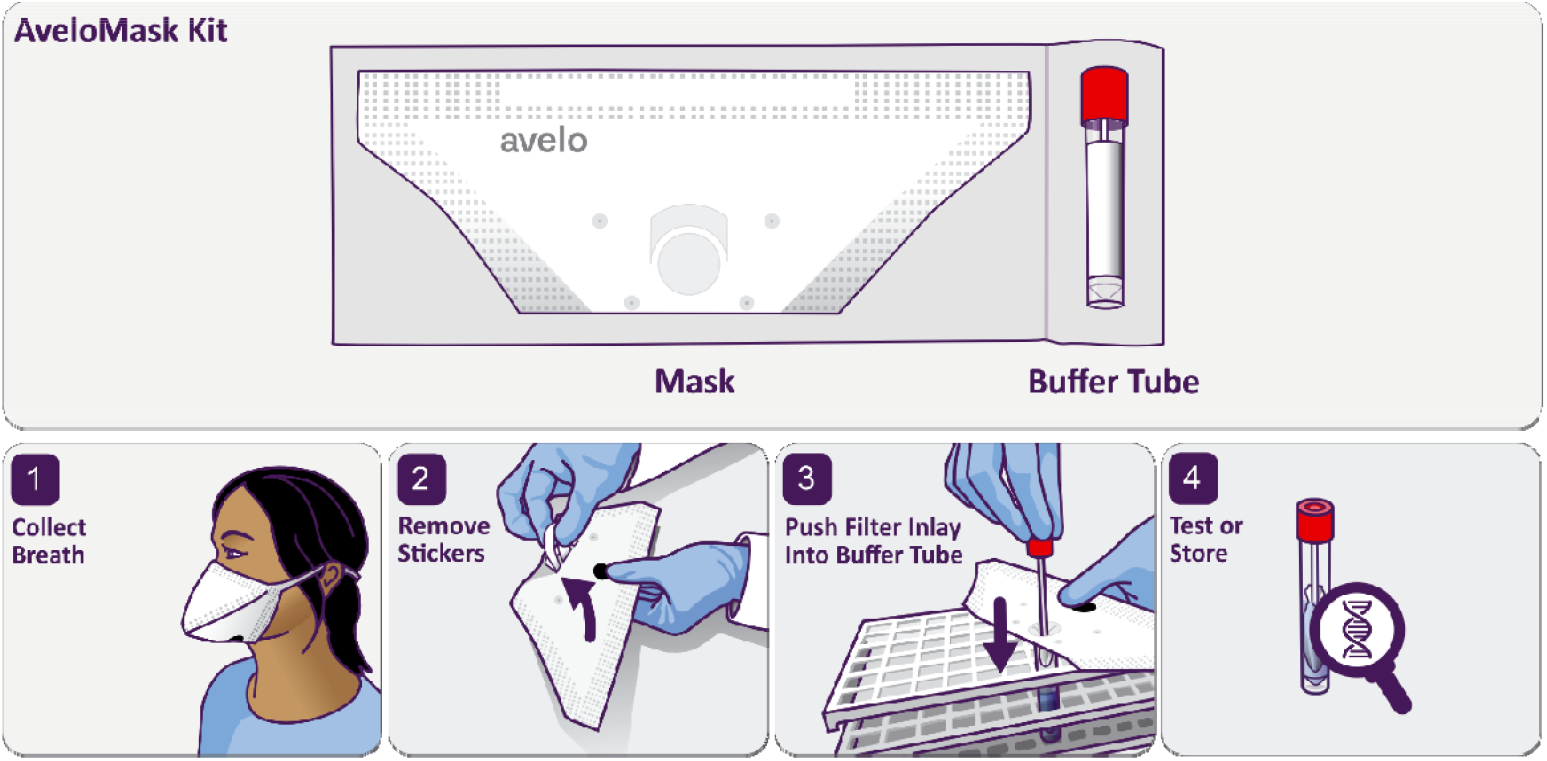
AveloMask Kit and breath aerosol collection procedure.

### Analytical evaluation

The aerosol filtration efficiency of the filter inlay was evaluated according to EN13274-7 using a PMFT 1000 test system (Palas, Germany). The assessment was conducted on 10 representative filter areas of 56 cm^2^ with paraffin oil aerosol particles generated by a PLG 1000 aerosol generator (Palas, Germany). Aerosol size distribution was measured using a Promo 1000 aerosol photometer (Palas, Germany).

The capture of Mycobacterial aerosols on the filter inlay was tested by exposing the filter to a nebulized bacterial suspension of *Mycobacterium bovis* BCG tagged with green fluorescent protein (BCG-GFP ^24^) as summarized in the supplemental material.

### Clinical evaluation

#### Study participants

To demonstrate the efficient capture and detection of MTB in exhaled breath aerosols, we enrolled n=61 participants between May 5 and October 17, 2024. We enrolled consenting adults (aged ≥18 years) with symptoms of TB at four outpatient primary health facilities in Kraaifontein, Scottsdene, Bloekombos, and Wallacedene (Cape Town, South Africa). Eligible participants had a persistent cough for at least two weeks and at least one additional symptom, such as hemoptysis, weight loss, fever, night sweats, malaise, contact with an active TB patient, chest pain, or loss of appetite. We excluded individuals currently receiving antimycobacterial treatment, those treated for TB in the past twelve months, and those unwilling to provide informed consent. In the initial phase, we enriched the study population by preferentially enrolling individuals with positive sputum Xpert Ultra results (n=20). Thereafter, we enrolled participants consecutively (independent from the Xpert Ultra results) from the same population.

#### Ethics and reporting

The Stellenbosch University Health Research Ethics Committee (HREC) approved the study (No. N16/07/089). Written informed consent was obtained from patients, as per the study protocols. Study participation did not affect the standard of care. This study is reported in accordance with the Standards for Reporting of Diagnostic Accuracy Studies guidelines (see STARD checklist in supplemental material).^25^

#### Sample collection

We collected detailed demographic and clinical data via standardized electronic case report forms (eCRFs) in a secure GCP/21 CFR part 11-compliant REDCap database.^26^ Sputum samples were obtained for reference standard testing with Xpert Ultra (Cepheid, USA) and MGIT culture after decontamination with 1% NaOH-NALC (Becton Dickinson, USA). The average interval between sputum collection and mask breath sampling was 1.8 days (range: 0–15 days). Participants wore the mask for 45 min, coughing deeply five times at the beginning and five times at the end of the collection period, in addition to any naturally occurring coughs.

Mask samples were processed on-site by trained community health workers. Filter inlays were transported in buffer at ambient temperature on the same day to the Stellenbosch University Biomedical Research Institute. There, they were stored at−20°C before being shipped on dry ice to Avelo, Switzerland, for blinded batch testing.

#### Usability assessment

We assessed the ease-of-use of the mask sampling procedure through structured questionnaires administered via eCRFs to both community health workers and participants. Usability questions used a standard 5-point Likert scale and were asked immediately after sample collection.

#### Mask sample processing and qPCR analysis

The frozen mask samples (consisting of filter inlays in stabilizing guanidinium thiocyanate buffer) were thawed, vortexed for 30 s, and placed in a water bath at 95°C for 1 h. After the heat elution/lysis, a disposable, sterile Pasteur pipette was used to compress the filter inlay and transfer the liquid into a new tube for subsequent extraction. Then, 1.5 ml of molecular-grade ethanol was added, and DNA was extracted from the entire sample using the QIAamp DNA Mini Kit (Qiagen, Germany), following the manufacturer’s instructions—omitting proteinase K and buffer AL, as the sample buffer already contains guanidinium thiocyanate. The samples were extracted and eluted with 100 µl Buffer AE. Every batch included a negative and positive process control (buffer and buffer spiked with 50 colony forming units BCG, respectively). Four 9 µl aliquots of DNA extract per sample were tested in 4 separate reactions. qPCR was performed using a QuantStudio5 96-well 0.2 ml thermal cycler (Thermo Fisher, Switzerland) with well-established primers and probes^27^ (see supplemental material for details). To assess the completeness of lysis, a subset of filter inlays from TB-positive participants underwent a second extraction. This was accomplished by adding 3 ml of guanidinium thiocyanate buffer and repeating the same processing and extraction protocol as described above. Mask assessors were blinded to clinical information and reference standard results.

#### Reference standard definitions

We used two reference standards: a composite microbiological reference standard (MRS) and a sputum Xpert MTB/RIF Ultra-based reference standard (SXRS). Under the MRS, participants were classified as having active TB if they had at least one positive sputum Xpert Ultra result and/or at least one positive sputum culture. Participants were considered MRS-negative if all sputum cultures and Xpert Ultra tests were negative, requiring a minimum of one negative sputum culture result. For the SXRS, participants were considered to have active TB if they had positive sputum Xpert Ultra results (very low or higher semiquantitative categories). Participants with a sputum Xpert Ultra trace-positive results were classified as indeterminate for the SXRS but considered in the MRS. For false positive mask results, participant records were reviewed in detail to extract information on chest X-ray, sputum smear microscopy, oral swab molecular results, antimycobacterial treatment, follow-up, and TB history, if available. Assessors of reference standard samples were blinded to index test results and clinical information.

#### Data analysis

A precision-based approach was used to determine the sample size, targeting two-sided 90% Wilson-score confidence intervals with a width of ±20%, assuming 65% sensitivity for mask qPCR. The assumed sensitivity, being lower than specificity and the study’s primary focus, guided the calculation, requiring 20 TB-positive participants.^28^ At least an equal number of TB-negative participants was enrolled.

We calculated the point estimates and 95% Wilson confidence intervals for the sensitivity and specificity of mask qPCR by comparison with the MRS and SXRS. Mean *IS6110* copy numbers from the four qPCR replicates of mask samples were used for ROC curve analysis by comparison with the SXRS using the pROC R package. In an exploratory subgroup analysis, sensitivity and mean *IS6110* copy numbers were assessed across sputum Xpert Ultra semiquantitative categories, MGIT culture time to positivity (TTP), and sex, to explore potential performance differences related to the typically smaller exhaled breath volume in female than in male participants. To assess the completeness of lysis, we determined the percent recovery of *IS6110* copy numbers from the first and second extractions of the same filter inlay, with the combined sum of copies from both extractions set to 100%. Analyses were performed via GraphPad Prism (version 10) and R (version 4.4.2).

## Results

### Analytical evaluation

Figure 2A shows the average fractional filtration efficiency of 10 filter inlays. The aerosol filtration efficiency of the filter inlay was greater than 80% for particles larger than 0.3 µm and efficiency was greater than 95% for particles larger than 0.5 µm. The cumulative filtration efficiency of 95.8% (95% CI: 94.5%−97.1%) for the particle size range of 0.3 to 2.2 µm suggests excellent capture of viral and bacterial aerosols by the filter inlay. Successful capture of bacterial aerosols was confirmed with nebulized BCG-GFP using microscopy (Figure 2B).

**Figure 2:**
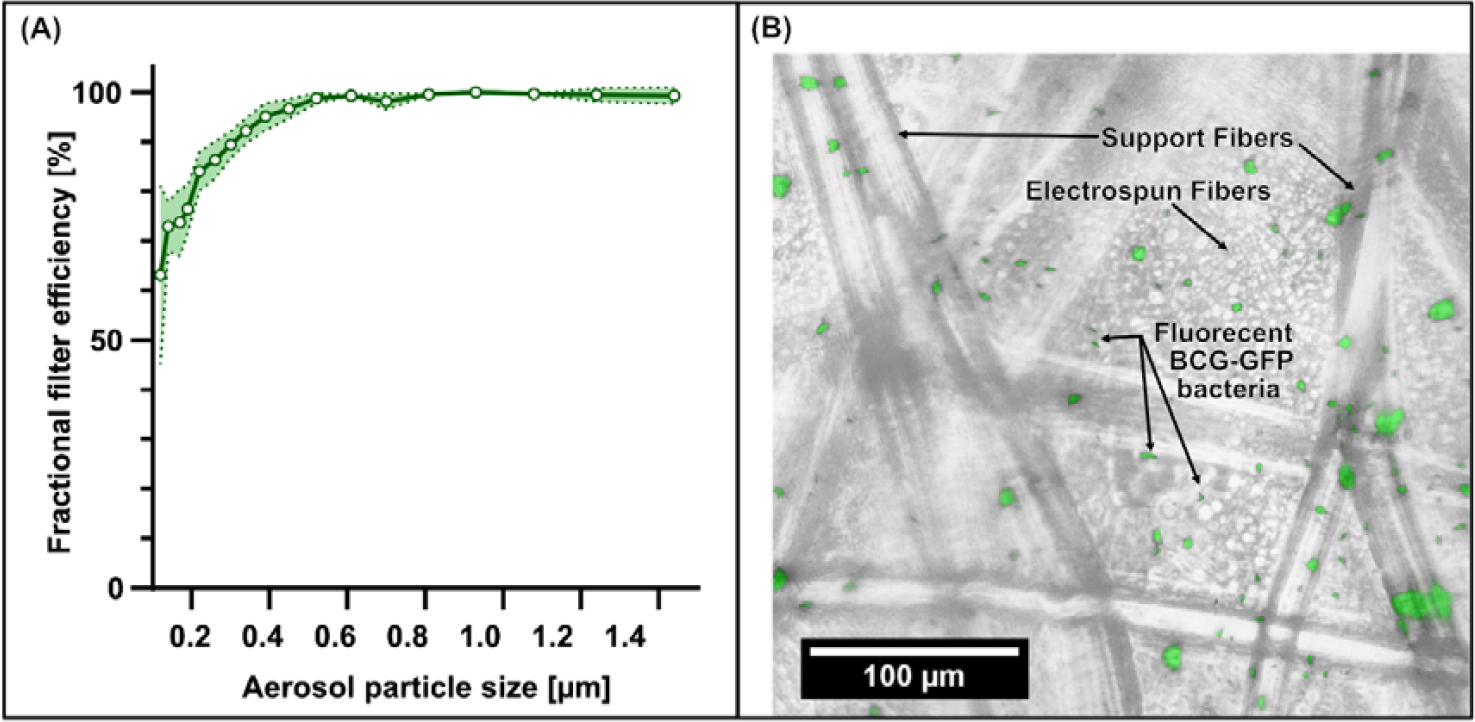
Filter inlay aerosol capture. (A) Fractional filtration efficiency (solid line) and 95% confidence intervals (dotted line) from 10 replicate tests of filter inlays showing high filtration efficiency for aerosol particle sizes above 0.3 µm, which is relevant for virus and bacteria. (B) Visualization of *Mycobacterium bovis* BCG tagged with green fluorescent protein (BCG-GFP) captured on the filter inlay. Green fluorescence channel image overlaid with a brightfield microscopy image showing rod-shaped BCG-GFP bacteria, indicating successful capture of nebulized mycobacterial aerosol.

### Clinical evaluation

#### Participant characteristics

Of the 61 enrolled participants, 3 (4.9%) were excluded from the main analysis because of filter processing errors during collection, leaving 58 participants for the main analysis (Figure 3). The participants primarily consisted of young adults, with an equal representation of both sexes. All participants exhibited symptoms suggestive of TB, with 43% (25/58) having an HIV infection, and 29% (17/58) having a history of prior TB. There was a good agreement between the MRS and SXRS with 59% (34/58) and 54% (31/57) of participants with confirmed TB respectively (Table 1).

**Table 1:** Demographic and clinical characteristics. Data are presented as median (interquartile range), or n (%).

| Characteristic | N=58 |
| --- | --- |
| Age | 41 (32, 46) |
| Sex at birth |  |
| Female | 31 (53%) |
| Male | 27 (47%) |
| Positive WHO tuberculosis symptom screen | 58 (100%) |
| Prior TB |  |
| Yes | 17 (29%) |
| No | 38 (66%) |
| Missing | 3 (5%) |
| People living with human immunodeficiency virus (HIV) |  |
| HIV Positive | 25 (43%) |
| HIV Negative | 29 (50%) |
| Missing | 4 (7%) |
| Microbiologic reference standard (MRS) |  |
| TB positive | 34 (59%) |
| TB negative | 24(41%) |
| Sputum Xpert Ultra reference standard (SXRS) |  |
| Positive | 31 (53%) |
| High | 8 (14%) |
| Medium | 9 (16%) |
| Low | 11 (19%) |
| Very Low | 3 (5%) |
| Negative | 26 (46%) |
| Indeterminate | 1 (1%) |
| Trace | 1 (1%) |

**Figure 3:**
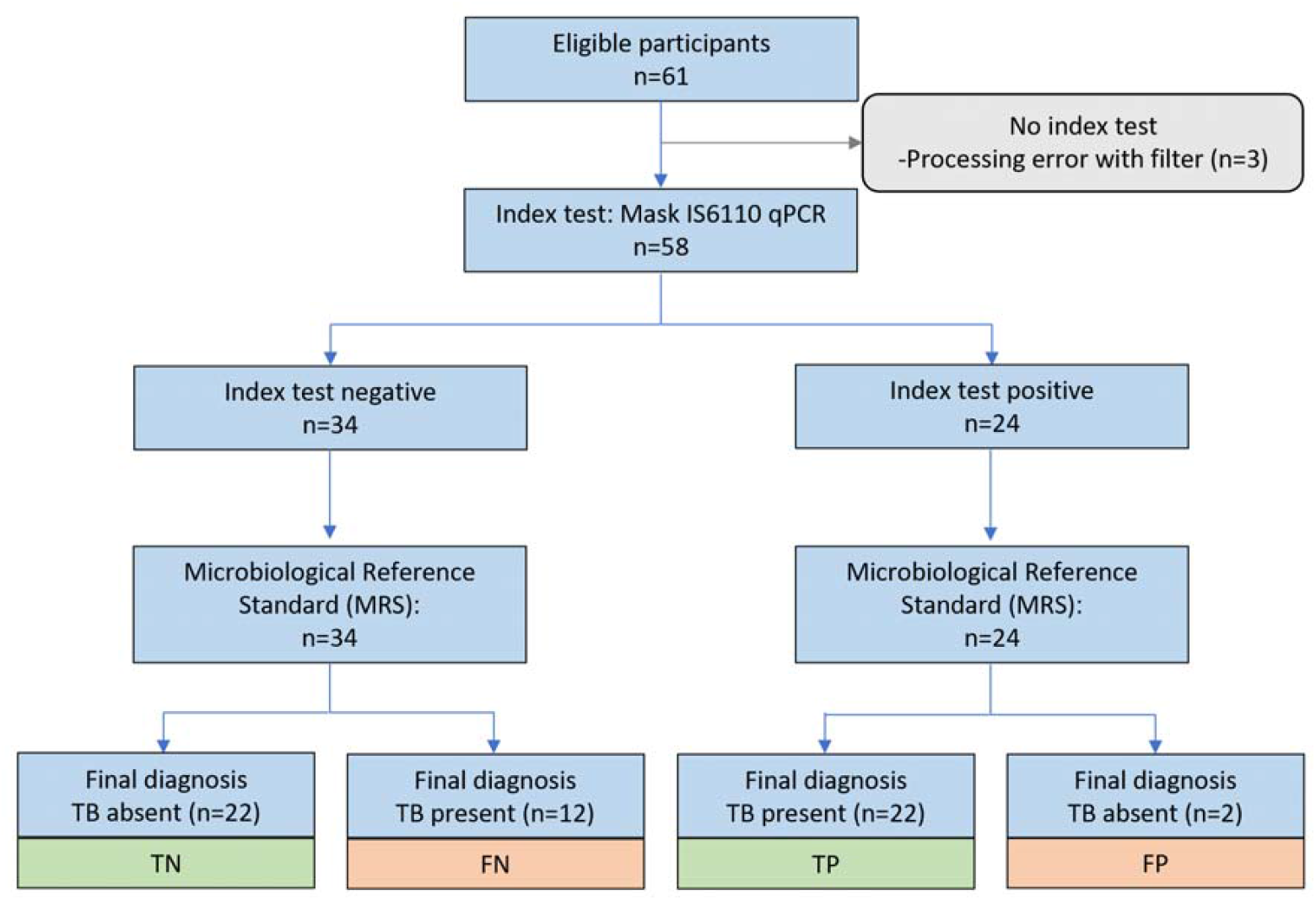
Study flow diagram.

#### Diagnostic accuracy of AveloMask qPCR testing

Among the 58 participants, concordance between the MRS and mask qPCR was 75.9% (95% CI: 63.5%–85.0%). Using the MRS, mask qPCR sensitivity was 64.7% (95% CI: 47.9%–78.5%) and specificity was 91.7% (95% CI: 74.2% –97.7%). When SXRS was used as the reference, mask qPCR sensitivity was 71.0% (95% CI: 53.4%– 83.9%) and specificity was 92.3% (95% CI: 75.9%–97.9%) (Figure 4). No signs of TB were found when the medical records of the two false positive participants were reviewed (see supplemental material). In both cases, positivity was observed in only 1 of 4 qPCR wells, with late cycle threshold (Ct) values (>38.5), suggesting very low positivity (Figure 5A). Negative extraction controls included in all qPCR runs remained negative, indicating minimal risk of cross-contamination.

**Figure 4:**
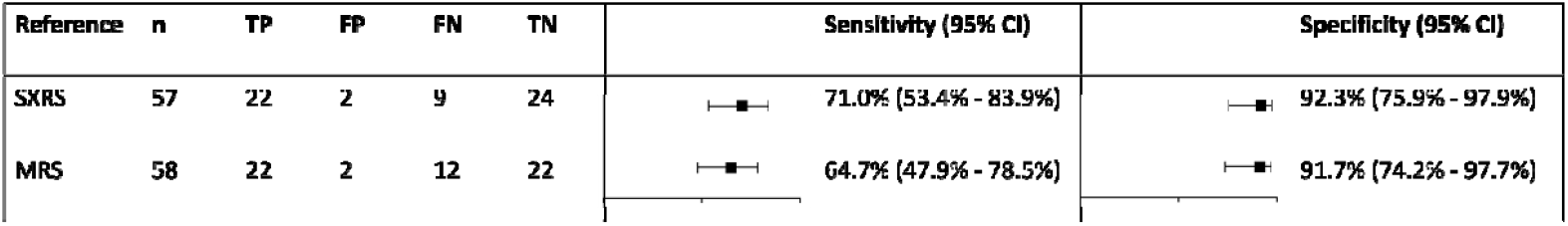
**Diagnostic accuracy of AveloMask qPCR** against the sputum Xpert Ultra reference standard (SXRS) and microbiological reference standard (MRS)

**Figure 5:**
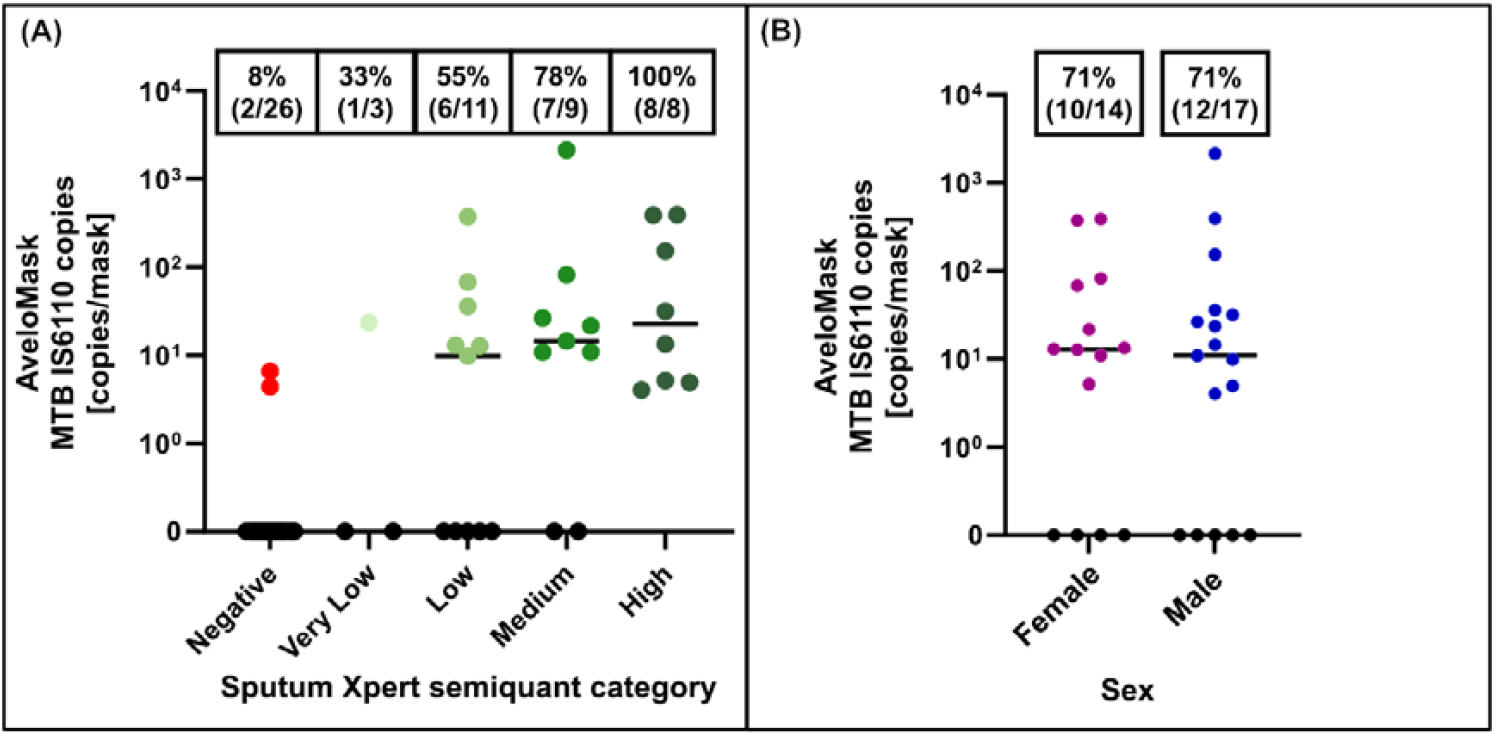
**Mean number of Mycobacterium tuberculosis IS6110 copies per AveloMask sample per participant** by (A) Xpert Ultra semiquantitative grade and (B) sex.

#### Quantification of MTB copy numbers from mask samples

Detectable levels of MTB *IS6110* copies were found in 71% (22/31) of mask samples collected from participants with SXRS-positive results. The IS6110 copy numbers spanned a 3-log range (4–2147 copies, mean: 175 copies) and showed a non-significant trend toward higher values in samples corresponding to higher sputum Xpert Ultra semiquantitative categories. Sensitivity increased with higher sputum Xpert Ultra semiquantitative categories, reaching 33%, 55%, 76%, and 100% for very low, low, medium, and high categories, respectively (Figure 5A). Mask positivity was higher among participants with high bacterial load, as indicated by shorter MGIT culture time to positivity (TTP): 100% (9/9) for samples with TTP ≤10 days compared to 36% (4/11) for samples with TTP >10 days. No significant differences in copy numbers or sensitivity were observed between sexes (Figure 5B). When using the mean *IS6110* copy numbers from mask qPCR for ROC curve analysis, sensitivity was 61.3% when optimized for 100% specificity (Figure S3 in supplemental material).

To assess DNA recovery, 19 filter inlays from TB-positive participants were reprocessed. There was a statistically significant trend towards higher *IS6110* copy numbers in the re-extracted samples (mean 62 vs. 113 copies; p=0.04), suggesting up to 65% of the DNA is missed in the initial extraction. All initially positive samples remained positive, and 3 of 5 of the initially false-negative samples were positive upon re-extraction (Figure S4 in supplemental material).

#### Usability Feedback

Usability feedback from all 61 participants indicated that the mask was generally well tolerated. A majority (90%) found the device at least “okay” to use, with 20% rating it as “very easy” and 34% rating it as “somewhat easy”. Only 10% of the participants reported the device as “somewhat difficult” to use, and none found it “very difficult” (Figure S2 in supplemental material). These findings suggest that the masks are user-friendly and feasible for use in primary care outpatient settings and among individuals presenting with symptoms of TB.

## Discussion

In this study of 61 individuals presenting with symptoms of TB, breath sampling with the novel AveloMask kit for 45 min followed by qPCR detection achieved a sensitivity of 71%, indicating effective capture of exhaled MTB. This sensitivity aligns closely with other face-mask sampling studies: Kodama et al.^21^ reported a sensitivity of 76% using 2-hour mask sampling coupled with a similar extraction method and loop-mediated isothermal amplification (LAMP), Williams et al.^20^ achieved an 86% sensitivity using qPCR following a 1-hour sampling period, and Shaikh et al.^23^ reported 75% sensitivity for a 10-min talk-cough-breathe process in children with PCR detection. However, unlike these previous methods, which require aseptic handling and filter manipulation with forceps, the AveloMask simplifies sample collection by enabling direct transfer of the filter inlay into an inactivating and nucleic acid-preserving buffer at the point of collection. Owing to its ease of use, the AveloMask kit demonstrated a successful collection rate of 95% (58/61) when administered by minimally trained community health workers in primary care outpatient settings.

Mask sampling was well tolerated by participants, making it an attractive alternative to sputum, which up to 18% of individuals are unable to produce and which is frequently of suboptimal quality.^3–5^ The high specimen availability for mask samples may offset its lower sensitivity, potentially diagnosing more individuals overall and achieving a higher diagnostic yield compared to molecular tests that require high-quality sputum specimens.^7,29^ Mask sampling should further be explored for the identification of asymptomatic individuals with subclinical TB that are estimated to be responsible for 68% of global transmission.^30^ >Since breath and cough aerosols are the primary modes of TB transmission, breath sampling may also help identify highly infectious individuals, who can be triaged for intervention strategies. Indeed, we observed higher mask qPCR positivity in individuals with high bacterial load sputum samples suggesting mask qPCR could be a measure for infectiousness.

Other mask sampling use cases worth researching include self-sampling, active case finding, and treatment monitoring—including in the context of vaccine and drug development. In a study by Fennelly and colleagues^31^, there was a rapid decrease in cough aerosol bacterial load within the first 3 weeks of effective treatment. Non-invasive breath sampling may also provide access to the lung microbiome ^32^, and improve the diagnosis of lower respiratory tract infections^33^ —which similarly suffer from limited availability of representative lower airway samples. Mask sampling may further serve as a valuable tool to study respiratory transmission of diseases^34,35^ and for pandemic preparedness as masks offer the dual benefit of protecting others from transmission while enabling diagnostic sampling from the wearer.

The success and widespread adoption of mask-based breath sampling will strongly depend on its compatibility with commercially available molecular testing platforms. Currently, *IS6110* copy numbers in mask samples are low, necessitating manual, column-based extraction of the entire sample. However, we observed incomplete DNA recovery, indicating that the actual bacterial load captured by masks is likely higher than currently measured. This suggests considerable potential to improve the elution, extraction, and lysis processes, the latter being particularly challenging due to the robust, thick cell wall of MTB. Given these considerations and supported by the high sensitivity reported by Kodama et al., who used a commercially available LAMP assay with a limit-of-detection around 100 CFU/ml, we anticipate that achieving high performance with commercial molecular platforms is feasible.^21^ Nevertheless, additional studies using optimized protocols are essential to realize this potential, in particular as the use of Xpert for mask samples has so far yielded insufficient sensitivity ranging from 13 to 48%.^23,36^

This study has several strengths. The AveloMask kit was carefully designed, produced, and analytically evaluated to demonstrate the capture efficiency of its novel fiber filter. It evaluates a non-invasive, breath-based sampling method that broadens TB diagnostics beyond sputum. Sample collection was conducted in real-world settings, and usability assessments confirmed feasibility in relevant populations with relevant users. Blinded qPCR testing, standardized protocols for sample handling and analysis, and the inclusion of process controls and standard curves ensured the reliability and reproducibility of results. Diagnostic accuracy was evaluated using two reference standards, including a composite MRS. The study population also included both people living with and without HIV, enhancing the relevance of findings.

While this study shows promising results supporting mask sampling as a non-sputum diagnostic alternative, it has several limitations. As a small proof-of-concept study, generalizability is limited and requires validation in larger studies, which are ongoing. Potential inclusion bias may have arisen from the case-control design and the restriction to sputum-producing participants. Future studies should follow the intention-to-diagnose principle and include those unable to expectorate to better assess diagnostic yield. ^7,37^ Two false positives could not be resolved, and despite precautions, procedural or environmental contamination remains a concern. *IS6110* copy number estimates should be interpreted with caution due to variability in extraction efficiency and qPCR stochasticity at low DNA levels. Current work is looking at optimizing the extraction protocol, which could further improve DNA recovery and thus sensitivity.

## Conclusion

The AveloMask breath aerosol sampling kit has shown promising diagnostic accuracy and feasibility in primary care settings, making it a valuable non-invasive diagnostic option for pulmonary TB. Given that at least one in five individuals struggle to produce sputum, breath-based sampling could significantly increase diagnostic yield, particularly among populations currently underserved by conventional diagnostics. Further optimization of DNA extraction methods and integration with commercial molecular testing platforms could enhance sensitivity and broaden clinical utility, ultimately contributing to earlier TB diagnosis and improved infection control.

## Data Availability

All data generated or analysed during this study are included in this published article, in the supplemental material, and supplemental data spreadsheet.

## List of abbreviations

BCG: *Mycobacterium bovis* Bacille Calmette–Guérin (Tuberculosis vaccine strain; member of the *Mycobacterium tuberculosis* complex)
CFU: Colony-forming units
CI: Confidence interval
Ct: Cycle threshold
DNA: Deoxyribonucleic acid
FMS: Face mask sampling
HIV: Human immunodeficiency virus
IS6110: Insertion sequence 6110 (MTB-specific DNA sequence)
LAMP: Loop-mediated isothermal amplification
MGIT: Mycobacterial Growth Indicator Tube
MRS: Microbiological reference standard
MTB: *Mycobacterium tuberculosis*
PCR: Polymerase chain reaction
qPCR: Quantitative polymerase chain reaction
ROC: Receiver operating characteristic
RSV: Respiratory syncytial virus
SDG: Sustainable Development Goal
SXRS: Sputum Xpert MTB/RIF Ultra reference standard
TB: Tuberculosis
VOC: Volatile organic compound
WHO: World Health Organization

## DECLARATIONS

### Ethics approval and consent to participate

Integrated in methods.

### Authors’ contributions

RV, KT, GT, and TB designed the study. ZB, JS, SMB, GT, and RV coordinated participant enrolment and clinical data collection. PR and CF produced mask kits and conducted aerosol capture experiments. PS provided BCG-GFP and designed *in vitro* experiments. HSK and RW analyzed clinical samples. KT performed statistical analysis and produced the video. TB and KT drafted the manuscript; all authors contributed, revised, and approved the final version. TB, RV, ZB, and KT had full data access, verified data integrity, and ensured accuracy of analyses. Authorship order follows contribution and ICMJE criteria.

## Acknowledgements

The authors thank the study participants and clinical staff, especially the Clinical Mycobacterial and Epidemiology clinical team for their contributions. We thank Theresa Heinrich (Blink Dx) for providing quantified IS6110 oligonucleotides. We thank Dr. Bettina Schulthess, Dr. Frank Imkamp and Dr. Tizian Griesser (Institute of Medical Microbiology, University of Zurich) for helpful discussion. ChatGPT was used for proofreading and text shortening; the authors carefully reviewed, edited, and assume full responsibility for the final content. Research in the laboratory of PS is supported by Swiss National Science Foundation and Federal Office of Public Health.

## Appendix

Supplemental Material (250408_Supplemental Material_v3.docx, figures, tables, supplementary methods)

Supplemental Data Spreadsheet (250408_Supplemental Data Spreadsheet_v1.xlsx, participant data)

Supplemental Movie (250403_Supplemental Movie_AveloMask Kit_Instructional_v3.mp4, Instructional Video)

## References

1. World Health Organization. 2024 Global Tuberculosis Report [Internet]. 2024 [cited 2025 Feb 28]. Available from: https:ww.who.int/teams/global-tuberculosis-programme/tb-reports/global-tuberculosis-report-2024

2. Silva S, Arinaminpathy N, Atun R, Goosby E, Reid M. Economic impact of tuberculosis mortality in 120 countries and the cost of not achieving the Sustainable Development Goals tuberculosis targets: a full-income analysis. Lancet Glob Health [Internet]. 2021 Oct;9(10):e1372–9. Available from: 10.1016/S2214-109X(21)00299-0

3. Papadopoulou P, Gaeddert M, Gupta-Wright A, Denkinger CM, Marx FM. Sputum availability and quality in country-level TB prevalence surveys. IJTLD OPEN [Internet]. 2024 Nov 1;1(11):528–30. Available from: https://www.ingentaconnect.com/content/10.5588/ijtldopen.24.0117

4. Kitonsa PJ, Sung J, Isooba D, Birabwa S, Naluyima I, Kakeeto J, et al. Quantifying sputum production success during community-based screening for TB. IJTLD OPEN [Internet]. 2024 Nov 1;1(11):522–4. Available from: http://www.ingentaconnect.com/content/10.5588/ijtldopen.24.0319

5. Broger T, Koeppel L, Huerga H, Miller P, Gupta-Wright A, Blanc FX, et al. Diagnostic yield of urine lipoarabinomannan and sputum tuberculosis tests in people living with HIV: a systematic review and meta-analysis of individual participant data. Lancet Glob Health [Internet]. 2023 Jun;11(6):e903–16. Available from: https://linkinghub.elsevier.com/retrieve/pii/S2214109x23001353

6. World Health Organization. Target product profiles for tuberculosis diagnosis and detection of drug resistance [Internet]. 2024 [cited 2025 Feb 28]. Available from: http://www.who.int/publications/i/item/9789240097698

7. Broger T, Marx FM, Theron G, Marais BJ, Nicol MP, Kerkhoff AD, et al. Diagnostic yield as an important metric for the evaluation of novel tuberculosis tests: rationale and guidance for future research. Lancet Glob Health [Internet]. 2024 Jul;12(7):e1184–91. Available from: http://www.ncbi.nlm.nih.gov/pubmed/38876764

8. Phillips M. Breath tests in medicine. Sci Am [Internet]. 1992 Jul;267(1):74–9. Available from: http://www.ncbi.nlm.nih.gov/pubmed/1502511

9. Chew N, Yun S, See KC. Diagnostic Accuracy of Breath Tests to Detect Pulmonary Tuberculosis: A Systematic Review. Lung [Internet]. 2025 Jan 22;203(1):26. Available from: http://www.ncbi.nlm.nih.gov/pubmed/39841224

10. Wang CC, Prather KA, Sznitman J, Jimenez JL, Lakdawala SS, Tufekci Z, et al. Airborne transmission of respiratory viruses. Science (1979) [Internet]. 2021 Aug 27;373(6558). Available from: https://www.science.org/doi/10.1126/science.abd9149;

11. Mubareka S, Groulx N, Savory E, Cutts T, Theriault S, Scott JA, et al. Bioaerosols and Transmission, a Diverse and Growing Community of Practice. Front Public Health [Internet]. 2019 Feb 21;7(FEB). Available from: https://www.frontiersin.org/article/10.3389/fpubh.2019.00023/full

12. Fennelly KP, Acuna-Villaorduna C, Jones-Lopez E, Lindsley WG, Milton DK. Microbial Aerosols: New Diagnostic Specimens for Pulmonary Infections. Vol. 157, Chest. Elsevier Inc; 2020. p. 540–6.

13. Liu Y, Ning Z, Chen Y, Guo M, Liu Y, Gali NK, et al. Aerodynamic analysis of SARS-CoV-2 in two Wuhan hospitals. Nature. 2020 Jun 25;582(7813):557–60.

14. Kim SH, Chang SY, Sung M, Park JH, Bin Kim H, Lee H, et al. Extensive Viable Middle East Respiratory Syndrome (MERS) Coronavirus Contamination in Air and Surrounding Environment in MERS Isolation Wards. Clinical Infectious Diseases [Internet]. 2016 Aug 1;63(3):363–9. Available from: https://academic.oup.com/cid/article-lookup/doi/10.1093/cid/ciw239

15. Kulkarni H, Smith CM, Lee DDH, Hirst RA, Easton AJ, O’Callaghan C. Evidence of Respiratory Syncytial Virus Spread by Aerosol. Time to Revisit Infection Control Strategies? Am J Respir Crit Care Med [Internet]. 2016 Aug;194(3):308–16. Available from: 10.1164/rccm.201509-1833OC

16. Yan J, Grantham M, Pantelic J, Bueno de Mesquita PJ, Albert B, Liu F, et al. Infectious virus in exhaled breath of symptomatic seasonal influenza cases from a college community. Proceedings of the National Academy of Sciences [Internet]. 2018 Jan 30;115(5):1081–6. Available from: 10.1073/pnas.1716561115

17. Patterson B, Dinkele R, Gessner S, Koch A, Hoosen Z, January V, et al. Aerosolization of viable Mycobacterium tuberculosis bacilli by tuberculosis clinic attendees independent of sputum-Xpert Ultra status. Proc Natl Acad Sci U S A. 2024 Mar 19;121(12).

18. Nduba V, Njagi LN, Murithi W, Mwongera Z, Byers J, Logioia G, et al. Mycobacterium tuberculosis cough aerosol culture status associates with host characteristics and inflammatory profiles. Nature Communications. 2024 Dec 1;15(1).

19. Theron G, Limberis J, Venter R, Smith L, Pietersen E, Esmail A, et al. Bacterial and host determinants of cough aerosol culture positivity in patients with drug-resistant versus drug-susceptible tuberculosis. Nat Med. 2020;26(9):1435–43.

20. Williams CM, Abdulwhhab M, Birring SS, De Kock E, Garton NJ, Townsend E, et al. Exhaled Mycobacterium tuberculosis output and detection of subclinical disease by face-mask sampling: prospective observational studies. Lancet Infect Dis [Internet]. 2020;20(5):607–17. Available from: 10.1016/S1473-3099(19)30707-8

21. Kodama T, Chikamatsu K, Kamada K, Mizuno K, Morishige Y, Igarashi Y, et al. Evaluation of testing face-mask filter samples with LAMP shows high rates of detection in pulmonary TB. The International Journal of Tuberculosis and Lung Disease [Internet]. 2024 Oct 1;28(10):476–81. Available from: http://www.ingentaconnect.com/content/10.5588/ijtld.24.0190

22. Williams CML, Cheah ESG, Malkin J, Patel H, Otu J, Mlaga K, et al. Face Mask Sampling for the Detection of Mycobacterium tuberculosis in Expelled Aerosols. Cardona PJ, editor. PLoS One [Internet]. 2014 Aug 14;9(8):e104921. Available from: https://dx.plos.org/10.1371/journal.pone.0104921

23. Shaikh A, Sriraman K, Vaswani S, Shah I, Poojari V, Oswal V, et al. SMaRT-PCR: sampling using masks and RT-PCR, a non-invasive diagnostic tool for paediatric pulmonary TB. The International Journal of Tuberculosis and Lung Disease [Internet]. 2024 Apr 1;28(4):189–94. Available from: http://www.ingentaconnect.com/content/10.5588/ijtld.23.0291

24. Dal Molin M, Selchow P, Schäfle D, Tschumi A, Ryckmans T, Laage-Witt S, et al. Identification of novel scaffolds targeting Mycobacterium tuberculosis. J Mol Med [Internet]. 2019 Nov 14;97(11):1601–13. Available from: http://link.springer.com/10.1007/s00109-019-01840-7

25. Bossuyt PM, Reitsma JB, Bruns DE, Gatsonis CA, Glasziou PP, Irwig L, et al. STARD 2015: an updated list of essential items for reporting diagnostic accuracy studies. BMJ [Internet]. 2015 Oct 28;351(12):1446–52. Available from: http://www.clinchem.org/cgi/doi/10.1373/clinchem.2015.246280

26. Harris PA, Taylor R, Minor BL, Elliott V, Fernandez M, O’Neal L, et al. The REDCap consortium: Building an international community of software platform partners. Vol. 95, Journal of Biomedical Informatics. Academic Press Inc.; 2019.

27. Chakravorty S, Simmons AM, Rowneki M, Parmar H, Cao Y, Ryan J, et al. The New Xpert MTB/RIF Ultra: Improving Detection of Mycobacterium tuberculosis and Resistance to Rifampin in an Assay Suitable for Point-of-Care Testing. Nacy CA, editor. mBio [Internet]. 2017 Sep 6;8(4):e00812–17. Available from: http://mbio.asm.org/lookup/doi/10.1128/mBio.00812-17

28. Drain PK, Gardiner JL, Hannah H, Broger T, Dheda K, Fielding K, et al. Guidance for Studies Evaluating the Accuracy of Biomarker-Based Nonsputum Tests to Diagnose Tuberculosis. J Infect Dis [Internet]. 2019;220(Supplement_3):S108–S115. Available from: http://fdslive.oup.com/www.oup.com/pdf/production_in_progress.pdf

29. Yerlikaya S, Broger T, Isaacs C, Bell D, Holtgrewe L, Gupta-Wright A, et al. Blazing the trail for innovative tuberculosis diagnostics. Infection [Internet]. 2023 Nov 30;(0123456789). Available from: 10.1007/s15010-023-02135-3

30. Emery JC, Dodd PJ, Banu S, Frascella B, Garden FL, Horton KC, et al. Estimating the contribution of subclinical tuberculosis disease to transmission: An individual patient data analysis from prevalence surveys. Elife [Internet]. 2023 Dec 18;12. Available from: https://elifesciences.org/articles/82469

31. Fennelly KP, Martyny JW, Fulton KE, Orme IM, Cave DM, Heifets LB. Cough-generated Aerosols of Mycobacterium tuberculosis. Am J Respir Crit Care Med [Internet]. 2004 Mar;169(5):604–9. Available from: http://www.atsjournals.org/doi/abs/10.1164/rccm.200308-1101OC

32. Chiyaka TL, Nyawo GR, Naidoo CC, Moodley S, Clemente JC, Malherbe ST, et al. PneumoniaCheck, a novel aerosol collection device, permits capture of airborne Mycobacterium tuberculosis and characterisation of the cough aeromicrobiome in people with tuberculosis. Ann Clin Microbiol Antimicrob [Internet]. 2024 Aug 22;23(1):74. Available from: https://ann-clinmicrob.biomedcentral.com/articles/10.1186/s12941-024-00735-x

33. Gal M, Francis NA, Hood K, Villacian J, Goossens H, Watkins A, et al. Matching diagnostics development to clinical need: Target product profile development for a point of care test for community-acquired lower respiratory tract infection. PLoS One. 2018 Aug 1;13(8).

34. Zhou J, Singanayagam A, Goonawardane N, Moshe M, Sweeney FP, Sukhova K, et al. Viral emissions into the air and environment after SARS-CoV-2 human challenge: a phase 1, open label, first-in-human study. Lancet Microbe. 2023 Aug 1;4(8):e579–90.

35. Hernaez B, Muñoz-Gómez A, Sanchiz A, Orviz E, Valls-Carbo A, Sagastagoitia I, et al. Monitoring monkeypox virus in saliva and air samples in Spain: a cross-sectional study. Lancet Microbe [Internet]. 2023 Jan;4(1):e21–8. Available from: https://linkinghub.elsevier.com/retrieve/pii/S2666524722002919

36. Hassane-Harouna S, Braet SM, Decroo T, Camara LM, Delamou A, Bock S De, et al. Face mask sampling (FMS) for tuberculosis shows lower diagnostic sensitivity than sputum sampling in Guinea. Ann Clin Microbiol Antimicrob [Internet]. 2023 Sep 7;22(1):81. Available from: https://ann-clinmicrob.biomedcentral.com/articles/10.1186/s12941-023-00633-8

37. Evans SR, Pennello G, Zhang S, Li Y, Wang Y, Cao Q, et al. Intention-to-diagnose and distinct research foci in diagnostic accuracy studies. Lancet Infect Dis [Internet]. 2025 Mar; Available from: https://linkinghub.elsevier.com/retrieve/pii/S1473309925000702

