## Supplemental Material for "AveloMask, a novel breath aerosol collection kit for airborne *Mycobacterium tuberculosis*: a proof-of-principle assessment"

**Table of Contents**

Figure S1: AveloMask Kit User Instructions	2

Supplemental Movie: AveloMask Kit instructions	3

Figure S2: Participant Usability Feedback	4

Figure S3: ROC curve based on IS6110 copies from AveloMask qPCR against sputum Xpert	5

Figure S4: Re-processing of filter inlays	6

Table S1: Filter Efficiency Raw Data	7

Additional Data for False Positive Participants	8

Supplementary Methods	9

STARD Checklist	11

### Figure S1: AveloMask Kit User Instructions

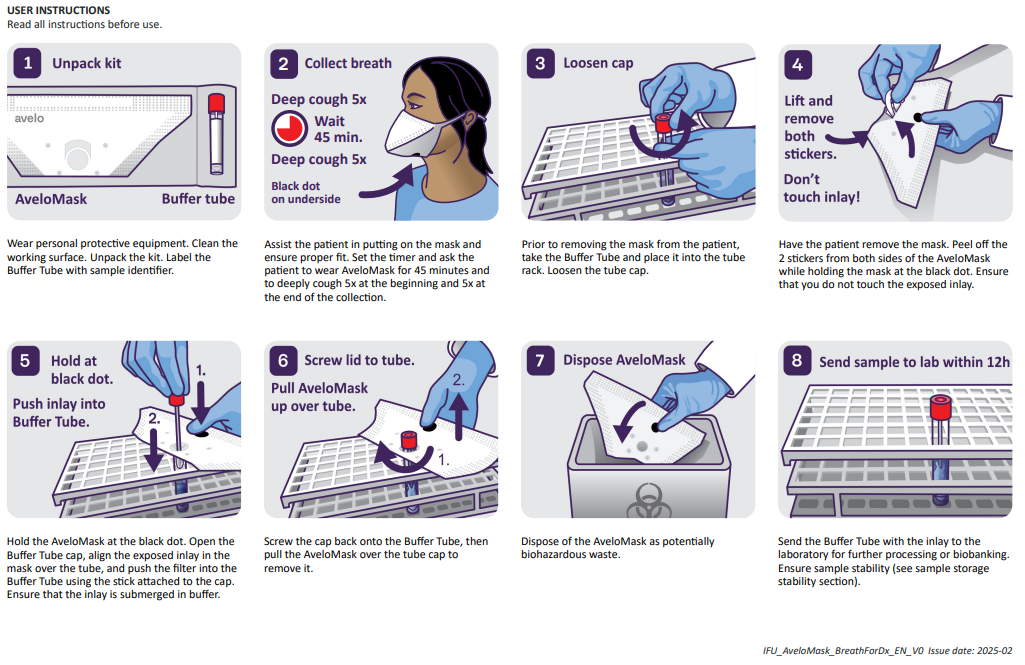

### Supplemental Movie: AveloMask Kit instructions

The movie shows the steps to collect and process an AveloMask sample

### Figure S2: Participant Usability Feedback

| **Patient Usability Feedback** | **N=61** |
| --- | --- |
| Ease-of-Use |  |
| Very Easy | 12 (20%) |
| Somewhat Easy | 21 (34%) |
| Okay | 22 (36%) |
| Somewhat Difficult | 6 (10%) |
| Very Difficult | 0 (0%) |

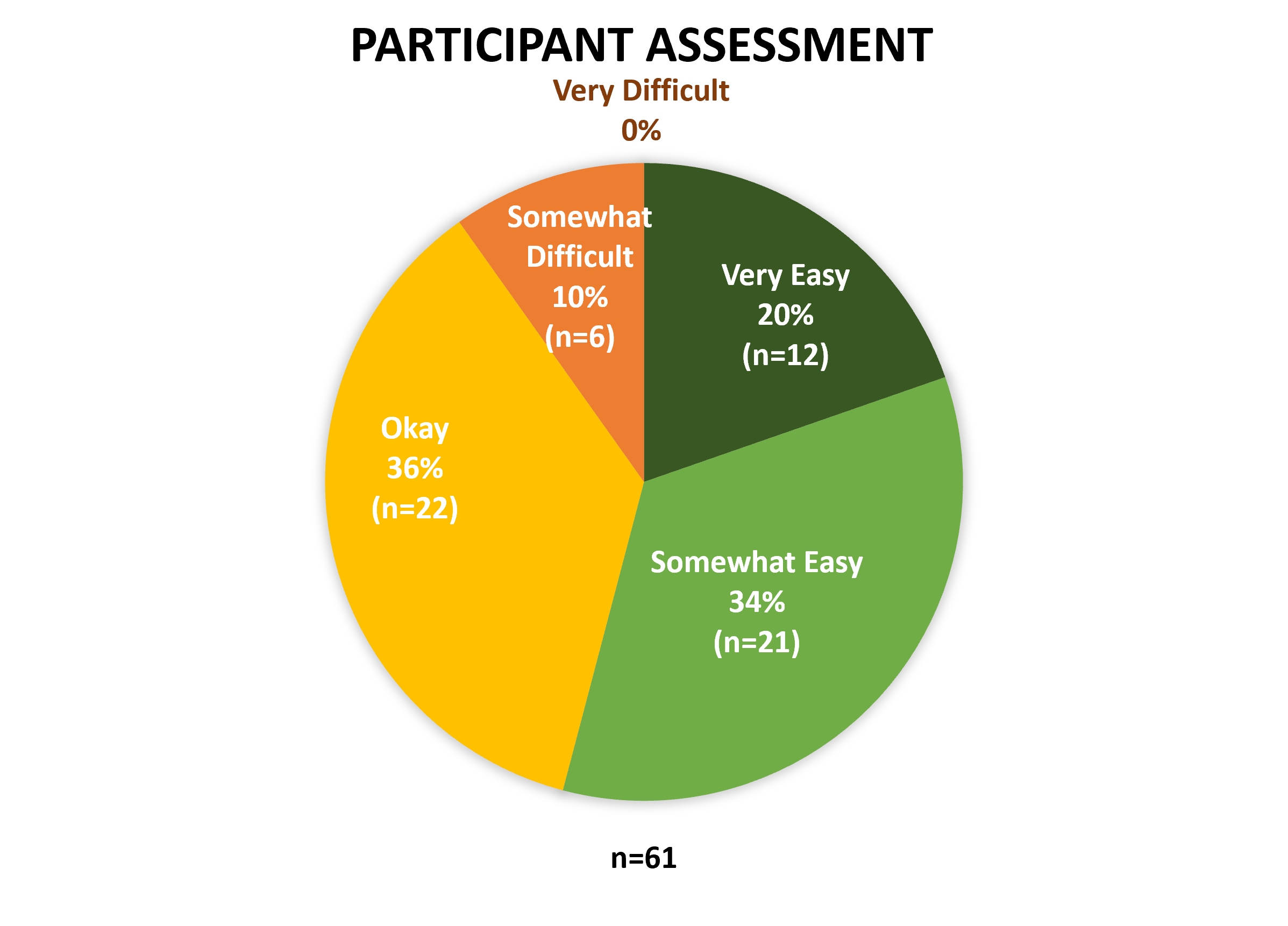

### Figure S3: ROC curve based on IS6110 copies from AveloMask qPCR against sputum Xpert

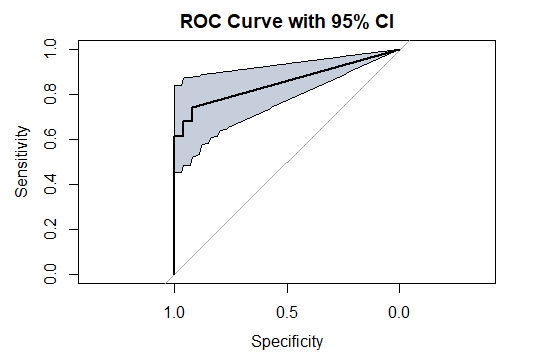

### Figure S4: Re-processing of filter inlays

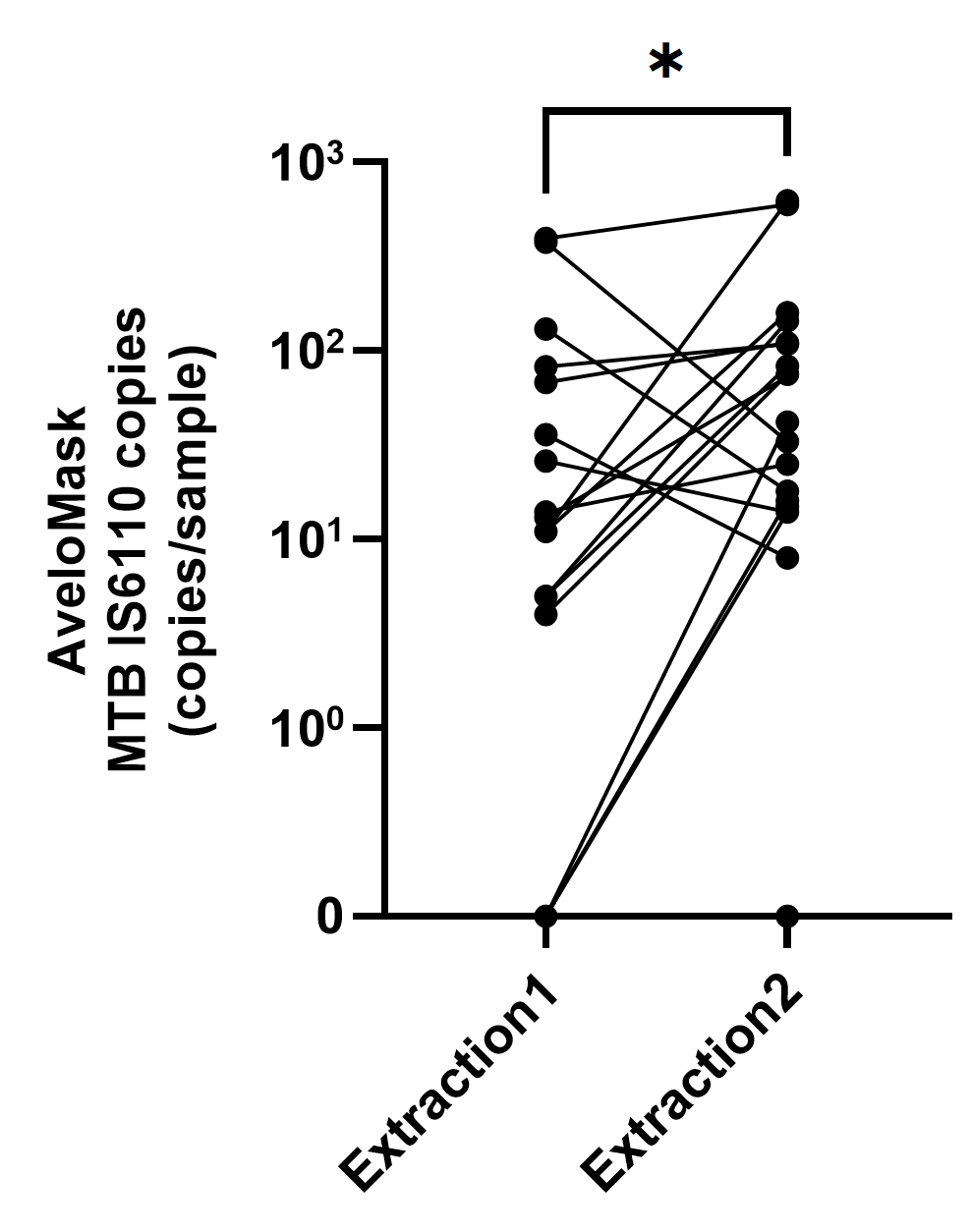

### Table S1: Filter Efficiency Raw Data

|  | Repeats - Identification | | | | | | | | | |
| --- | --- | --- | --- | --- | --- | --- | --- | --- | --- | --- |
| Particles size  [um] | P503 | P505 | P507 | P509 | P552 | P554 | P556 | P558 | P586 | P588 |
| 0.124 | 70% | 64% | 49% | 64% | 0% | 84% | 84% | 71% | 60% | 85% |
| 0.144 | 66% | 73% | 72% | 68% | 88% | 70% | 81% | 68% | 67% | 76% |
| 0.166 | 72% | 74% | 71% | 76% | 88% | 61% | 80% | 87% | 62% | 66% |
| 0.192 | 76% | 71% | 79% | 77% | 83% | 79% | 80% | 86% | 64% | 69% |
| 0.221 | 82% | 84% | 86% | 80% | 94% | 81% | 88% | 90% | 76% | 79% |
| 0.255 | 83% | 88% | 87% | 90% | 93% | 90% | 90% | 87% | 77% | 78% |
| 0.295 | 85% | 87% | 88% | 86% | 92% | 94% | 97% | 89% | 89% | 87% |
| 0.341 | 89% | 92% | 95% | 93% | 98% | 96% | 90% | 90% | 86% | 93% |
| 0.393 | 90% | 91% | 98% | 94% | 100% | 98% | 96% | 97% | 89% | 98% |
| 0.454 | 94% | 96% | 97% | 96% | 100% | 100% | 98% | 98% | 96% | 91% |
| 0.525 | 97% | 98% | 98% | 98% | 100% | 100% | 100% | 100% | 96% | 100% |
| 0.606 | 98% | 100% | 98% | 99% | 100% | 100% | 100% | 100% | 98% | 100% |
| 0.700 | 95% | 98% | 96% | 94% | 100% | 100% | 100% | 100% | 98% | 100% |
| 0.808 | 100% | 97% | 100% | 100% | 100% | 100% | 100% | 100% | 98% | 100% |
| 0.933 | 100% | 100% | 100% | 100% | 100% | 100% | 100% | 100% | 100% | 100% |
| 1.077 | 100% | 100% | 96% | 100% | 100% | 100% | 100% | 100% | 100% | 100% |
| 1.244 | 100% | 100% | 94% | 100% | 100% | 100% | 100% | 100% | 100% | 100% |
| 1.437 | 100% | 100% | 93% | 100% | 100% | 100% | 100% | 100% | 100% | 100% |

### Additional Data for False Positive Participants

**AVL074-R2D2031247 Clinical Data Summary**

Participant had initial visit 11/10/2024:

- Chest X-Ray: Abnormal
  - No cavitation present
  - Classification as “TB Possible“
- Gave 3 sputum samples, all mucosalivary in quality
  - Sputum Xpert: Negative
  - Sputum smear (2x): Negative
  - Sputum MGIT (2x): Negative
  - Sputum swab tested with NAAT: Negative
- Gave 1 tongue swab
  - Tongue swab tested with NAAT: Negative
- Participant was not started on TB treatment and is considered TB negative

Participant did not return for a follow-up visit

**AVL068-R2D2031253 Clinical Data Summary**

Participant had initial visit 10/10/2024:

- Chest X-Ray: Normal
- Gave 3 sputum samples, all salivary in quality
  - Sputum Xpert: Negative
  - Sputum smear (2x): Negative
  - Sputum MGIT (2x): Negative
  - Sputum swab tested with Pluslife: Negative
- Gave 1 tongue swab
  - Tongue swab tested with Pluslife: Negative
- Participant was not started on TB treatment and is considered TB negative

Participant had a 3-month follow up visit 17/01/2025:

- Participant did not report any symptoms at follow-up visit
- Follow-up Chest X-ray: Normal
- Participant gave sputum sample at follow-up visit (quality not specified)
  - Sputum Xpert: Negative
- Patient is still considered TB negative

### Supplementary Methods

**AveloMask Kit Production**

Filter inlays were produced via electrospinning using a Fluidnatek LE-100 system (Bioinicia Fluidnatek SLU, Spain). A 12% (w/v) solution of Polyamide 6 (Mw = 55,600 Da, BASF, Switzerland) in acetic acid/formic acid (2:1 w/w, from Carl Roth, Switzerland, and ABCR, Switzerland, respectively) was prepared and electrospun onto a spunbond polypropylene substrate following a proprietary protocol. The resulting fiber mats were cut into 75 mm × 75 mm sheets and heat-sealed along the edges using a custom-made stamping and sealing machine to form the filter inlay. The filter inlay is subsequently mounted in a releasable manner on the inner side of a custom-made duckbill face mask. Prior to inserting the filter inlay, two custom-made peel-off stickers were applied on each side of the face mask to allow processing of the filter inlay after breath collection as described in the user instructions above. Buffer tubes were produced by attaching a polyamide stick to the tube lid of standard lab tubes (Copan, Italy) and filling tubes with 3 mL of guanidinium thiocyanate buffer.

**Aerosol Testing with nebulized BCG-GFP**

Filter inlays capture of Mycobacterial aerosols on the filter inlay was tested by exposing the filter to a nebulized bacterial suspension of *Mycobacterium bovis* BCG tagged with green fluorescent protein (BCG-GFP). Approx. 80 µl of bacterial suspension containing ~700,000 colony forming units (cfu) BCG-GFP was nebulized using a Cirrus2 nebulizer (Intersurgical, UK) at an air mass flow of 6 l/min for 15 s and diluted with 34 l/min filtered air. The resulting bioaerosol was directed into a custom-built housing containing a circular filter disk with a diameter of 24 mm. Air mass flow rates were controlled using two SFC5500 flow controllers and measured using an SFM3300 flow sensor (Sensirion, Switzerland) with custom-made LabView software (National Instruments, Switzerland). See picture of the setup below. After aerosol collection, the filter disks were removed and microscopically observed with a Biotek Cytation 5 cell imaging reader (Agilent, Switzerland). Phase contrast images and fluorescent images of the same field-of-view were overlaid using Affinity Photo software (Serif Ltd., UK).

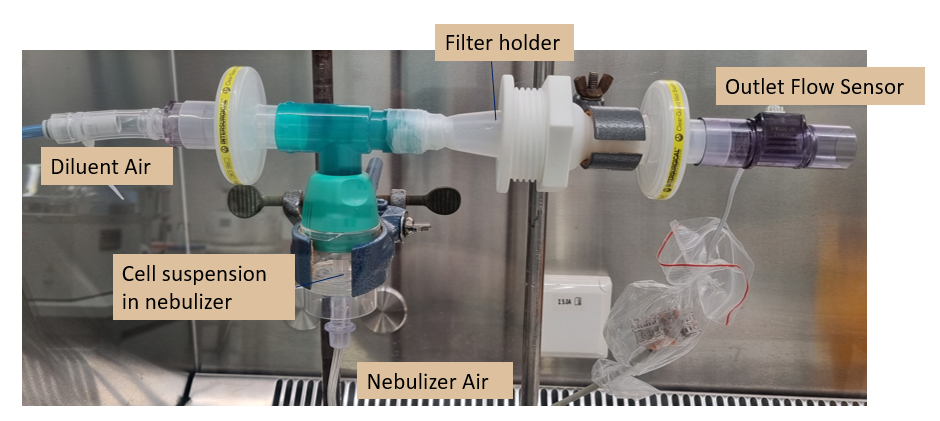

**PCR analysis of Mask samples**

Each reaction contained 9 µl DNA extract, 10 µl master mix (PrimeTime Gene Expression Master Mix, IDT, Switzerland), 1 µl Primers and Probe mix (final concentration in PCR reaction: 1000 nM each primer and 250 nM probe). The primers and probe targeting the MTB-specific IS6110 insertion sequence were based on Chakrovorty et al. 2017 with forward primer (5'-AGCGCCGCTTCGGACCACCAG-3'), reverse primer (5'-AGGCGTCGGTGACAAAGGCCACGTA-3'), and probe (5'-CY5-CGGCTGTGGGTAGCAGACCTCACC-IBRQ-3'). The amplification protocol consisted of an initial activation at 95°C for 3 min, followed by 55 cycles of denaturation at 95°C for 15 s, and annealing/extension at 66°C for 60 s. In addition to the negative and positive process controls, a five-level standard curve (ranging from 10 to 5000 copies) was generated using a synthesized oligonucleotide of the IS6110 target sequence. This standard curve was included in each run and used to estimate IS6110 copy numbers in the samples. The copy number for the control material was determined using digital PCR (QX200 system, Biorad, Germany).

### STARD Checklist

|  | **Section & Topic** | **No** | **Item** | **Done?** |
| --- | --- | --- | --- | --- |
|  | **TITLE OR ABSTRACT** |  |  |  |
|  |  | **1** | Identification as a study of diagnostic accuracy using at least one measure of accuracy  (such as sensitivity, specificity, predictive values, or AUC) | yes |
|  | **ABSTRACT** |  |  |  |
|  |  | **2** | Structured summary of study design, methods, results, and conclusions  (for specific guidance, see STARD for Abstracts) | yes |
|  | **INTRODUCTION** |  |  |  |
|  |  | **3** | Scientific and clinical background, including the intended use and clinical role of the index test | yes |
|  |  | **4** | Study objectives and hypotheses | yes |
|  | **METHODS** |  |  |  |
|  | *Study design* | **5** | Whether data collection was planned before the index test and reference standard  were performed (prospective study) or after (retrospective study) | yes |
|  | *Participants* | **6** | Eligibility criteria | yes |
|  |  | **7** | On what basis potentially eligible participants were identified  (such as symptoms, results from previous tests, inclusion in registry) | yes |
|  |  | **8** | Where and when potentially eligible participants were identified (setting, location and dates) | yes |
|  |  | **9** | Whether participants formed a consecutive, random or convenience series | yes |
|  | *Test methods* | **10a** | Index test, in sufficient detail to allow replication | yes |
|  |  | **10b** | Reference standard, in sufficient detail to allow replication | yes |
|  |  | **11** | Rationale for choosing the reference standard (if alternatives exist) | NA |
|  |  | **12a** | Definition of and rationale for test positivity cut-offs or result categories  of the index test, distinguishing pre-specified from exploratory | yes |
|  |  | **12b** | Definition of and rationale for test positivity cut-offs or result categories  of the reference standard, distinguishing pre-specified from exploratory | yes |
|  |  | **13a** | Whether clinical information and reference standard results were available  to the performers/readers of the index test | yes |
|  |  | **13b** | Whether clinical information and index test results were available  to the assessors of the reference standard | yes |
|  | *Analysis* | **14** | Methods for estimating or comparing measures of diagnostic accuracy | yes |
|  |  | **15** | How indeterminate index test or reference standard results were handled | yes |
|  |  | **16** | How missing data on the index test and reference standard were handled | yes |
|  |  | **17** | Any analyses of variability in diagnostic accuracy, distinguishing pre-specified from exploratory | yes |
|  |  | **18** | Intended sample size and how it was determined | yes |
|  | **RESULTS** |  |  |  |
|  | *Participants* | **19** | Flow of participants, using a diagram | yes |
|  |  | **20** | Baseline demographic and clinical characteristics of participants | yes |
|  |  | **21a** | Distribution of severity of disease in those with the target condition | yes |
|  |  | **21b** | Distribution of alternative diagnoses in those without the target condition | no |
|  |  | **22** | Time interval and any clinical interventions between index test and reference standard | yes |
|  | *Test results* | **23** | Cross tabulation of the index test results (or their distribution)  by the results of the reference standard | yes |
|  |  | **24** | Estimates of diagnostic accuracy and their precision (such as 95% confidence intervals) | yes |
|  |  | **25** | Any adverse events from performing the index test or the reference standard | yes |
|  | **DISCUSSION** |  |  |  |
|  |  | **26** | Study limitations, including sources of potential bias, statistical uncertainty, and generalisability | yes |
|  |  | **27** | Implications for practice, including the intended use and clinical role of the index test | yes |
|  | **OTHER INFORMATION** |  |  |  |
|  |  | **28** | Registration number and name of registry | no |
|  |  | **29** | Where the full study protocol can be accessed | no |
|  |  | **30** | Sources of funding and other support; role of funders | Yes |
